# The clinical outcomes of acute kidney injury substages based on serum cystatin C in pediatric patients undergoing cardiac surgery

**DOI:** 10.1101/2023.04.25.23289121

**Authors:** Yinan Li, Dongyun Bie, Chao Xiong, Sheng Shi, Zhongrong Fang, Zhongyuan Lu, Jianhui Wang

## Abstract

**Background:** Multiple biomarkers have been identified by previous studies to diagnose acute kidney injury (AKI). Moreover, combination of biomarkers with conventional criteria to define AKI substages so that we can identify high-risk patients and improve diagnostic accuracy were recommended. Our study aimed to explore the incidence of AKI substages defined by serum cystatin C (CysC), determine whether AKI substages were associated with worse outcomes.

**Methods:** We prospectively included 2519 pediatric patients (<16yrs) undergoing cardiac surgery with cardiopulmonary bypass in our cohort between March 2022 to February 2023 in Fuwai Hospital. Demographic and clinical variables we collected. To define AKI substages, Kidney Disease: Improving Global Outcomes AKI definition (based on SCr or CysC) was used. The association between AKI exposure and outcomes including length of intensive care unit stay, duration of mechanical ventilation, length of hospital stay and 30-day mortality was assessed. In addition, we determined areas under the receiver operating characteristic curve and cutoff value of CysC at different timepoints to predict AKI.

**Results:** 507 (20.8%) patients developed SCr-AKI, with 337 (13.8%) in stage 1, 77(3.2%) in stage 2 and 93 (3.8%) in stage 3 respectively. Of the 1925 patients without SCr-AKI, 256 (14.3%) met the definition of sub-AKI. Of the 507 patients with SCr-AKI, 281 (55.4%) patients were defined as AKI substage A, while others (226, 44.6%) were defined as AKI substage B. After adjusting for BSA, neonates, STAT mortality score≥4, previous sternotomy and CPB time>120min, the postoperative LOIS, LOHS and DMV were prolonged with increasing hospitalization expense (P<0.05) in patients with SCr-AKI and/or CysC-AKI. Meanwhile, only the hospitalization expense was increased in patients with SCr-AKI(P<0.05) after the same adjustment. The AUC was 0.691, 0.720 and 0.817 respectively in ROC curves of preoperative, relative variation of or postoperative serum CysC. Delong’ test showed that postoperative serum CysC might have better diagnostic performance characteristic than preoperative or relative variation of CysC (P<0.001), with a cutoff point at 1.29 mg/dL (Specificity, 0.77; Sensitivity, 0.71)

**Conclusions:** Our analysis indicates defining AKI with both CysC and SCr might more significantly affecting clinical outcome associations in pediatric patients undergoing cardiac surgery. Moreover, the serum CysC cutoff of 1.29mg/dL postoperatively is a valuable threshold for AKI risk assessment to define AKI subtypes.

## Background

Acute kidney injury (AKI) occurs frequently in pediatric cardiac surgery, which is significantly associated with increased mortality and morbidities, prolonged length of hospital stay, and much more medical expense^1-3^. The reported incidences of AKI after pediatric cardiac surgery ranged from 27% to 52%^3-5^, while management options for AKI are limited. The common supportive strategies include maintaining stable hemodynamic, avoiding nephrotoxic agents, prudent fluid balance and renal replacement therapy. Therefore, the early diagnosis for AKI plays important roles in maintaining patients’ safety.

The international consensus conference of Acute Dialysis Quality Initiative (ADQI) recommends the combination of damage and functional biomarkers, along with clinical information in identifying high-risk patient groups and improve the diagnostic accuracy of AKI^6^. Previous studies have defined AKI substages A and B by the absence or presence of biomarkers that might varied due to the absence of oliguria ^7-9^. However, whether isolated elevation in SCr or biomarkers was associated with worse outcomes in pediatric patients undergoing cardiac surgery remained elusive^10-12^. Moreover, the threshold of biomarkers obtained in adults is difficult to directly use in pediatric population, thus leaving the threshold varying a lot by ages and weights in this population^13^.

Cystatin C (CysC) is a low-molecular-weight protein generated endogenously by all nuclear cells and freely filtered by the glomerulus, which is more highly associated with glomerular filtration rate (GFR) than SCr^14^. Thus, the increased level of serum CysC might directly reflect renal injury. Meanwhile, previous studies demonstrated that serum CysC are not affected by age, gender, muscle mass, disease or maternal creatinine^15^. Several authors have demonstrated that CysC can be used an early biomarker of AKI in adults and children^12,16,17^. However, evidence for the serum CysC use in diagnosing AKI in pediatric patients undergoing cardiac surgery with cardiopulmonary bypass (CPB). Furthermore, the optimal threshold is urgently necessary to be explored.

Therefore, the aims of our cohort study were to explored the incidence of substage AKI, the threshold of CysC in pediatric patients undergoing cardiac surgery, in conjunction with classical SCr criteria^18^, and to determine whether AKI substage were associated with adverse outcomes in this population.

## Materials and Methods

### Study Population

Our study was conducted between March 2022 to February 2023 in Fuwai Hospital. All pediatric patients (<16yrs) undergoing cardiac surgery with cardiopulmonary bypass were included in our prospective cohort. Exclusion criteria included gestational age less than 34 weeks, weight less than 2kg at time of surgery, end-stage kidney disease, by history, estimated glomerular filtration rate less than 25mL/min/1.73m^2^ within 3 months before surgery, or most recent preoperative SCr value of more than 3.5mg/dL, preoperative dialysis, a history of renal transplant and unavailable preoperative SCr, CysC and height data. The study was approved by the Ethical Committee of Fuwai Hospital (Approval No. 2021-1607) and registered at ClinicalTrials (https://clinicaltrials.gov/) (NCT05489263, A predictive Score System for AKI Following Pediatric Cardiac Surgery). Written informed consent was obtained from patients’ legal representatives before enrollment.

### Data collection

Demographic and clinical variables we collected included age, body surface area (BSA), gender, history of previous sternotomy, preoperative use of loop diuretics, CPB time, aortic cross-clamping (ACC) time, intraoperative use of corticosteroids and postoperative renal replacement therapy. Society of Thoracic Surgeons-European Association for Cardio-Thoracic Surgery (STAT) mortality score contains 5 categories differentiating surgical mortality risk in accordance with surgery procedures^19^. Height was recorded to calculate preoperative SCr-estimated GFR (eGFR) according to the modified Schwartz formula: eGFR=0.413*height (cm)/SCr (mg/dL) ^20^.

### Definition of AKI

The Kidney Disease: Improving Global Outcome (KDIGO) criteria for SCr was applied^21^. The routinely measured SCr most recent within 7 days preoperatively and every 24 hours within postoperative day (POD) 7 were used to identify SCr-AKI. Stage 1 is at least a 50% SCr rise from baseline within 7 days or a 0.3mg/dL rise within 48 hours, stage 2 is SCr doubling, and stage 3 is SCr tripling requiring dialysis or an eGFR of less than 35 mL/min/1.73 m^2^ at any time.

### AKI substages

Patients were identified with non-AKI, sub-AKI, AKI substage A, and AKI substage B based on the combination of CysC and SCr. The CysC criteria was defined by applying the Kidney Disease: Improving Global Outcomes definition but using CysC change instead of SCr change similar to what has been done by others (ie. definitions of stage 1, stage 2, and stage 3 above) ^12,17^.

### Clinical Outcomes

Postoperative length of intensive care unit stay (LOIS), length of hospital stay (LOHS), duration of mechanical ventilation (DMV), hospitalization expense and 30-day mortality were assessed as clinical outcomes.

### Statistical Analysis

Continuous variables were expressed as mean (standard deviation) if they were considered to be normally distributed after the Shapiro–Wilk test, or else expressed as median (interquartile range). Categorical variables are expressed as numbers (proportions). For comparison across groups, continuous variable were performed using orthogonal contrasts in analysis of variance, and categorical variables were compared using χ 2 test or Fisher exact test. Comparisons were made across the following 4 SCr-AKI and CysC-AKI definition combinations: no AKI by SCr or CysC (Non-AKI), SCr-AKI only (AKI substage A), CysC-AKI only (Subclinical AKI), and AKI by SCr and CysC (AKI substage B). Percentage agreement and κ statistic were calculated to evaluate agreement between CysC-AKI and SCr-AKI definitions (presence or absence of AKI and classification across AKI severity stages). Logistic regression was used to acquire the adjusted association between SCr-AKI or CysC-AKI and 30-day mortality. Univariable and multivariable Poisson regression was used to evaluate associations of SCr-AKI and CysC-AKI with LOIS, LOHS, DMV and hospitalization expense, controlling for BSA, neonates, STAT mortality score≥4, previous sternotomy, and CPB time>120 minutes. The selection of variables was based on previous study and our clinical experience^12^. The areas under the receiver operating characteristic curve (ROC) and 95% CIs of preoperative CysC, CysC variation or postoperative CysC to predict AKI were also calculated, difference between any 2 of the ROCs was tested by deLong’s test. The sensitivity, specificity and cut-off point were also derived from ROCs. All tests related to ROC were performed using r package “pROC”. STATA version 14.0 (Stata Corporation, College Station, TX) and R version 4.2.1 (R Foundation for Statistical Computing) were used for data analysis.

## Results

### Patients and AKI substage

In the study period, 2519 patients were recruited, 87 of whom were excluded due to unavailable preoperative SCr, CysC, and height data, known congenital abnormality of kidney and failure to collect postoperative SCr and CysC. Therefore, the finial cohort eligible for analyzing included 2432 patients (Figure 1). As was shown in Figure 1, 507 (20.8%) patients developed SCr-AKI, with 337 (13.8%) in stage 1, 77(3.2%) in stage 2 and 93 (3.8%) in stage 3 respectively. Furthermore, when additionally classifying patients into different substage basing on SCr and CysC, the incidence of non-AKI, sub-AKI, and AKI substages A or B in each stage of SCr-AKI were displayed in Figure 1. Of the 1925 patients without SCr-AKI, 256 (14.3%) patients had a serum CysC increase higher than 50% from the baseline CysC, thus meeting the definition of sub-AKI. Of the 507 patients with SCr-AKI, 281 (55.4%) patients with a serum CysC increase no more than 50% from the baseline CysC were defined as AKI substage A, while others (226, 44.6%) were defined as AKI substage B.

**Figure 1.**
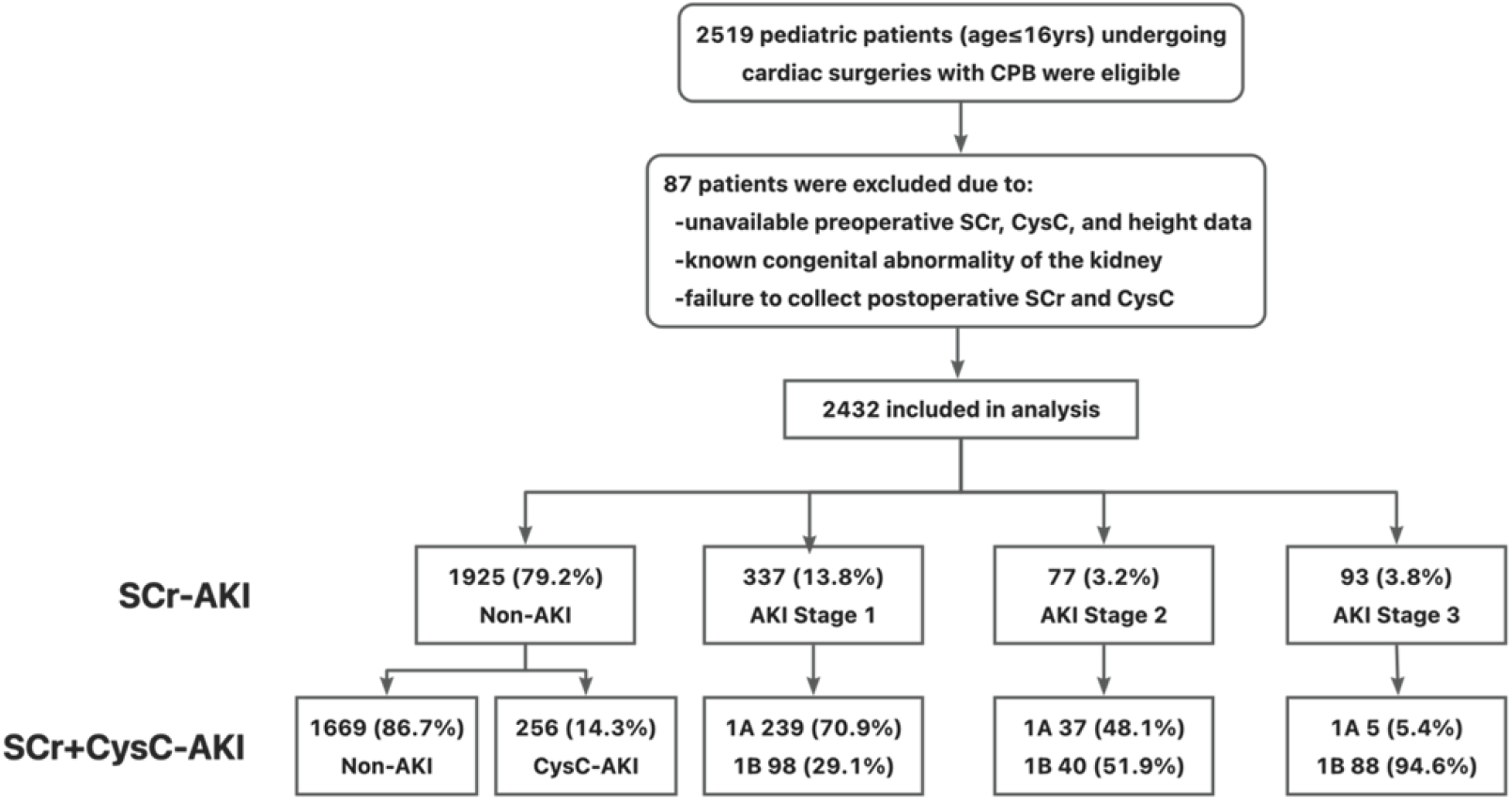
Flow diagram demonstrating study design Abbreviations: CysC, cystatin C; SCr-AKI, serum creatinine-defined AKI; CPB, cardiopulmonary bypass.

Demographic and clinical data grouped by 4 types of AKI were displayed in Table 1. Patients with AKI substage B had the worst risk profile for AKI comparing with patients with AKI substage A, including older age, lower preoperative SCr-eGFR, more neonates, higher incidence of previous sternotomy, longer CPB and ACC time. Patients with AKI substage B also had the longest postoperative LOIS, LOHS, or DMV and hospitalization expense across all AKI definition methods, tending to a more severe SCr-AKI severity pattern (increased proportion of stage 2 and 3 SCr-AKI) than patients with only SCr-AKI. The intraoperative corticosteroids were comparable between the four groups. 7 patients were treated with extracorporeal membrane oxygenation (ECMO) and 17 patients were given renal replacement therapy (RRT).

**Table 1.** Patients’ characteristics and outcomes stratified by subgroups of AKI

|  | Non-AKI n=1925 |  | AKI n=507 |  | P value |
| --- | --- | --- | --- | --- | --- |
|  | Non-AKI<br>n=1669 | Subclinical AKI<br>n=256 | AKI substage A<br>n=281 | AKI substage B<br>226 |  |
| **<u>Patients' characteristics</u>** |  |  |  |  |  |
| Age, mean (SD), d | 1498 (1284) | 1673 (1346) | 484 (580) | 631 (1058)<sup>a</sup> | <0.001 |
| BSA, m<sup>2</sup> | 0.68 (0.32) | 0.72 (0.32) | 0.42 (0.17) | 0.41 (0.28) | <0.001 |
| Gender, male, No. (%) | 850 (50.9) | 149 (58.2) | 147 (52.3) | 139 (61.5) | 0.006 |
| Neonate, No. (%) | 17 (1.0) | 0 (0) | 9 (3.2) | 52 (23)<sup>a</sup> | <0.001 |
| Preoperative SCr-estimated GFR, median (IQR), mL/min/1.73 m<sup>2</sup> | 122.6<br>(97.3,143.6) | 128.1<br>(105.1,145.2) | 119.9<br>(98.8,146.6) | 112.6 (69.7,138.6)<br><sup>a</sup> | <0.001 |
| Preoperative CysC, median (IQR), mg/L | 0.91 (0.80,1.06) | 0.78 (0.67, 0.90) | 1.04 (0.92,1.23) | 1.05 (0.86,1.39) | <0.001 |
| Previous sternotomy, No. (%) | 152 (9.1) | 38 (14.8) | 15 (5.3) | 24 (10.6)<sup>a</sup> | 0.002 |
| Preoperative use of loop diuretic, No. (%) | 129 (7.7) | 33 (12.9) | 42 (15.0) | 61 (27.0)<sup>a</sup> | <0.001 |
| STAT mortality score, No. (%) |  |  |  |  | <0.001 |
| 1 | 667 (40.0) | 86 (33.6) | 91 (32.4) | 45 (19.9) |  |
| 2 | 746 (44.7) | 113 (44.1) | 139 (49.5) | 78 (34.5) |  |
| 3 | 117 (7.0) | 29 (11.3) | 16 (5.7) | 43 (19.0) |  |
| 4 | 136 (8.1) | 26 (10.2) | 34 (12.1) | 58 (25.6) |  |
| 5 | 3 (0.2) | 2 (0.8) | 1 (0.3) | 2 (0.9) |  |
| CPB time, median (IQR), min | 72 (48,106) | 89 (57,136) | 83 (55,122) | 121 (83,155)<sup>a</sup> | <0.001 |
| ACC time, median (IQR), min | 43 (27,71) | 56 (34,92) | 50 (32,81) | 71 (50,106)<sup>a</sup> | <0.001 |
| Intraoperative corticosteroids, No. (%) | 1309 (78.4) | 211 (82.4) | 225 (80.1) | 179 (79.2) | 0.52 |
| **<u>Outcomes</u>** |  |  |  |  |  |
| SCr-AKI, No. (%) |  |  |  |  | <0.001 |
| No AKI | 1669 (86.7) | 256 (14.3) | 0 (0) | 0 (0) |  |
| Stage 1 | 0 (0) | 0 (0) | 239 (70.9) | 98 (29.1) |  |
| Stage 2 | 0 (0) | 0 (0) | 37 (48.1) | 40 (51.9) |  |
| Stage 3 | 0 (0) | 0 (0) | 5 (5.4) | 88(94.6) |  |
| 30-day mortality, No. (%) | 11 (0.7) | 0 (0) | 3 (1.1) | 8 (3.5)<sup>a</sup> | 0.001 |
| Postoperative length, median (IQR), d |  |  |  |  |  |
| ICU stay | 1 (1,3) | 3 (2,5) | 3 (1,5) | 7 (3,15)<sup>a</sup> | <0.001 |
| Hospital stay | 6 (5,9) | 7 (6,11) | 7 (5,11) | 12 (8,18)<sup>a</sup> | <0.001 |
| Mechanical ventilation | 0.17 (0.13,0.38) | 0.29 (0.17,1.00) | 0.38 (0.17,1.08) | 2.94 (0.75,7.33)<sup>a</sup> | <0.001 |
| Hospitalization expense, ×10000CNY | 6.65 (5.59,9.27) | 8.39 (6.32,12.32) | 8.41 (6.13,11.82) | 13.78 (9.64, 1.83)<sup>a</sup> | <0.001 |

Abbreviations: ACC, aortic cross-clamping time; AKI, acute kidney injury; BSA, body surface area; CPB, cardiopulmonary bypass; CysC, cystatin C; GFR, glomerular filtration rate; ICU, intensive care unit; IQR, interquartile range; SCr-AKI, serum creatinine-defined AKI; STAT mortality score, Society of Thoracic Surgeons-European Association for Cardio-Thoracic Surgery mortality score.
Statistically significant difference across the 4 groups was defined as $P < 0.001$ .
<sup>a</sup> Statistically significant difference between SCr-AKI only group vs AKI by SCr and CysC group ( $P < .005$ , correction for multiple comparisons).

As summarized in Table 2 and Table 3, AKI incidences defined by any SCr criteria or any CysC criteria were 547 of 2432 [22.5%] vs 482 of 2432 [19.8%]). Percentage agreement between any SCr-AKI and CysC-AKI was 76% (κ = 0.295, 95%CI, 0.279-0.303). There was 95% agreement (κ = 0.596, 95%CI, 0.529-0.663) between SCr-AKI and CysC-AKI stage 2 or worse. Percentage agreement between definitions for staging AKI severity was 77% (κ=0.296, 95%CI, 0.279-0.303). The highest discrepancy was in stage 1 AKI classification: of 337 children with stage1 SCr-AKI, 239 were classified as having no CysC-AKI.

**Table 2.** Comparison of AKI severity by CysC-AKI staging vs. SCr-AKI staging

| SCr-AKI staging | CysC-AKI staging |  |  |  | Total |
| --- | --- | --- | --- | --- | --- |
|  | Non-AKI | 1 | 2 | 3 |  |
| Non-AKI | 1669<sup>a</sup> | 232 | 23 | 1 | 1925 |
| 1 | 239 | 81<sup>a</sup> | 17 | 0 | 337 |
| 2 | 37 | 31 | 9<sup>a</sup> | 0 | 77 |
| 3 | 5 | 2 | 2 | 84<sup>a</sup> | 93 |
| Total | 1950 | 346 | 51 | 85 | 2432 |

Abbreviations: CysC-AKI, cystatin C-defined AKI; SCr-AKI, serum creatinine-defined AKI.
<sup>a</sup> Patients for whom there is agreement in AKI staging between the 2 definitions.
<sup>b</sup> kappa statistic (95%CI) for AKI severity staging is 0.295 (0.279-0.303) ( $P<0.001$ )

**Table 3.** Comparison of AKI stage ≥ 2 by CysC-AKI staging vs. SCr-AKI staging

| SCr-AKI Stage 2 Staging | CysC-AKI Stage 2 Staging |  |  |
| --- | --- | --- | --- |
|  | No AKI or Stage 1 | Stage 2 or 3 | Total |
| No AKI or Stage 1 | 2221<sup>a</sup> | 41 | 2262 |
| Stage 2 or 3 | 75 | 95<sup>a</sup> | 170 |
| Total | 2296 | 136 | 2432 |

Abbreviations: CysC-AKI, cystatin C-defined AKI; SCr-AKI, serum creatinine-defined AKI.
a Patients for whom there is agreement in AKI staging between the 2 definitions.
b kappa statistic (95%CI) for AKI severity staging is 0.596 (0.529-0.663) ( $P<0.001$ )

### Clinical outcomes

Table 1 demonstrated the 30-day mortality of AKI substages. The 30-day mortality was higher in children with AKI substage B than that in those with non-AKI and other types of AKI, respectively. Multivariate logistic regression was used to adjust for BSA, STAT mortality score≥4 and CPB time>120min. The adjusted hazard ratio (HR) for 30-day mortality in patients with SCr-AKI was 1.59 (95%CI, 0.40-6.36, *P*=0.513), while that in patients with SCr-AKI and/or CysC-AKI was 1.34 (95%CI, 0.26-6.78, *P*=0.724) (Table 4). However, as was shown in Table 5, the postoperative LOIS, LOHS and DMV were prolonged with increasing hospitalization expense (*P*<0.05) in patients with SCr-AKI and/or CysC-AKI, after adjusted for BSA, neonates, STAT mortality score≥4, previous sternotomy and CPB time>120min. Meanwhile, only the hospitalization expense was increased in patients with SCr-AKI(*P*<0.05) after the same adjustment.

**Table 4.** Adjusted hazard ratio of 30-day moratlity in patients with SCr-AKI and/or CysC-AKI

|  | Hazard ratio | 95%CI | *P* value |
| --- | --- | --- | --- |
| SCr-AKI | 1.59 | 0.40-6.36 | 0.513 |
| SCr-AKI and/or CysC-AKI | 1.34 | 0.26-6.78 | 0.724 |

Model was adjusted for BSA, STAT mortality score ≥ 4, CPB time>120min.
Abbreviations: CysC-AKI, cystatin C-defined AKI; SCr-AKI, serum creatinine-defined AKI; BSA, body surface area; STAT mortality score, Society of Thoracic Surgeons-European Association for Cardio-Thoracic Surgery mortality score; CPB, cardiopulmonary bypass.

**Table 5.** Adjusted outcomes in patients with SCr-AKI and/or CysC-AKI

|  | Coefficient | Standard error | Beta | R-square | *P* value |
| --- | --- | --- | --- | --- | --- |
| Postoperative LOIS (d) |  |  |  |  |  |
| SCr-AKI | 0.26 | 0.25 | 0.02 | 0.30 | 0.316 |
| SCr-AKI and/or CysC-AKI | 0.77 | 0.28 | 0.06 | 0.33 | 0.013* |
| Postoperative LOHS (d) |  |  |  |  |  |
| SCr-AKI | 0.56 | 0.33 | 0.03 | 0.25 | 0.096 |
| SCr-AKI and CysC-only-AKI | 0.89 | 0.37 | 0.05 | 0.25 | 0.016* |
| Postoperative MVT (d) |  |  |  |  |  |
| SCr-AKI | 0.24 | 0.19 | 0.03 | 0.26 | 0.206 |
| SCr-AKI and CysC-only-AKI | 0.73 | 0.30 | 0.08 | 0.31 | 0.011* |
| Hospitalization expense (× 10000CNY) |  |  |  |  |  |
| SCr-AKI | 0.59 | 0.26 | 0.04 | 0.47 | 0.023* |
| SCr-AKI and CysC-only-AKI | 1.23 | 0.23 | 0.11 | 0.49 | <0.001* |

Model was adjusted for BSA, neonates, STAT mortality score ≥ 4, previous sternotomy, CPB time $>$ 120min.
Abbreviations: CysC-AKI, cystatin C-defined AKI; SCr-AKI, serum creatinine-defined AKI; BSA, body surface area; STAT mortality score, Society of Thoracic
Surgeons-European Association for Cardio-Thoracic Surgery mortality score; CPB, cardiopulmonary bypass.
\* $P<0.05$

### Diagnostic performance characteristics of CysC in AKI

ROC curves of preoperative, relative variation of or postoperative serum CysC, for AKI diagnosed with KDIGO criteria were shown in Figure 2. Table 6 presents the sensitivity, specificity, AUC and cutoff point of serum CysC at different time points and its relative variation. The AUC was 0.691, 0.720 and 0.817 respectively in ROC curves of preoperative, relative variation of or postoperative serum CysC. Delong’ test showed that postoperative serum CysC might have better diagnostic performance characteristic than preoperative or relative variation of CysC (*P*<0.001), with a cutoff point at 1.29 mg/dL (Specificity, 0.77; Sensitivity, 0.71). The cutoff points for preoperative and relative variation of serum CysC were 0.96 mg/dL (Specificity, 0.66; Sensitivity, 0.63) and 1.25-fold (Specificity, 0.63; Sensitivity, 0.70) separately.

**Table 6.** Characteristics of ROC curves for different types of CysC in diagnosing SCr-AKI

|  | AUC | Cutoff Value | Specificity | Sensitivity | *P* value |
| --- | --- | --- | --- | --- | --- |
| Preoperative CysC | 0.691<sup>c</sup> | 0.96 | 0.66 | 0.63 | <0.001 |
| Relative variation of CysC | 0.720<sup>c</sup> | 1.25 | 0.63 | 0.70 | <0.001 |
| Postoperative CysC | 0.817<sup>a,b</sup> | 1.29 | 0.77 | 0.71 | <0.001 |

<sup>a</sup>. *P* value < 0.001 in Delong's test between ROC curve for preoperative CysC and others
<sup>b</sup>. *P* value < 0.001 in Delong's test between ROC curve for Relative variation of CysC and others
<sup>c</sup>. *P* value < 0.001 in Delong's test between ROC curve for Postoperative CysC and others

**Figure 2.**
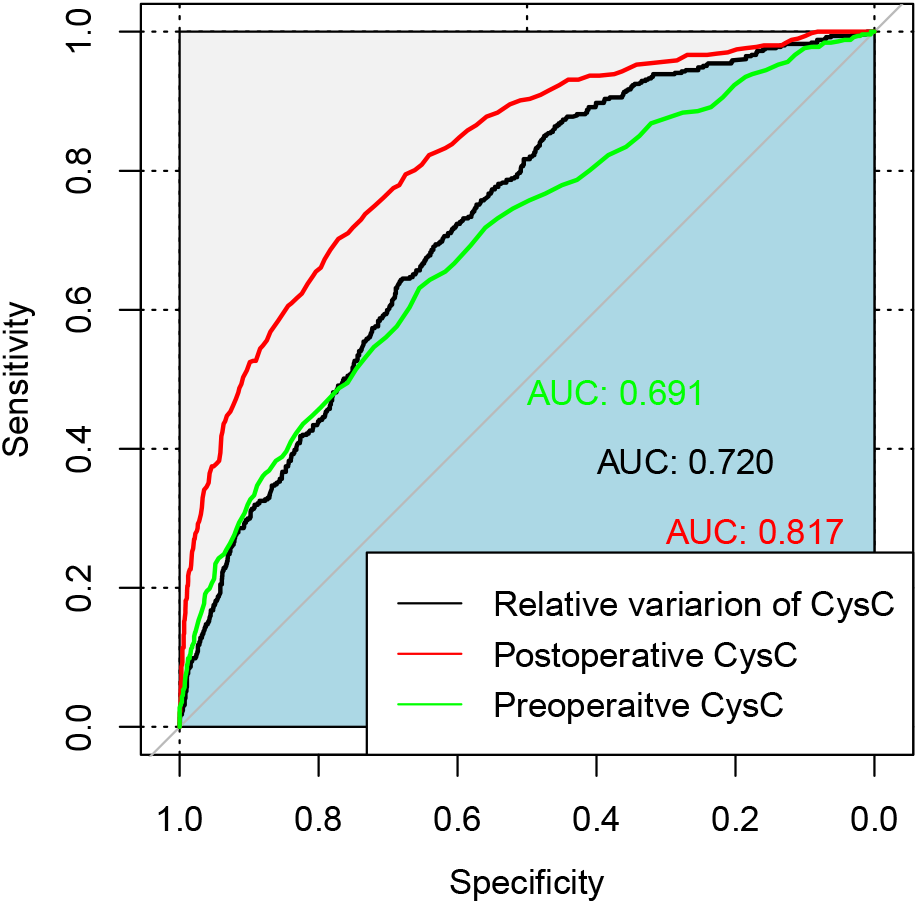
ROC curves for different types of CysC in diagnosing SCr-AKI Abbreviations: CysC, cystatin C; ROC curves, receiver operating characteristic curves; SCr-AKI, serum creatinine-defined AKI

## Discussion

Previous study has shown that CysC was an acceptable prognostic for AKI in pediatric patients, which might be more accurate in estimating GFR than SCr due to it is at constant production and independent of age, gender, muscle mass, disease or maternal creatinine. Previous study has shown CysC was more strongly associated with urine kidney injury biomarkers in children undergoing cardiac surgery^12^and much less affected by volume status than SCr^22^. However, the direct association and diagnostic performance of CysC for AKI was concerning till now. Moreover, the outcomes of patients defined as substage AKI by CysC remained inaccurate. We hypothesized that due to its small variation across different physical status and personnel, CysC might be a superior way to define AKI in clinical trials and outcome studies of AKI.

In our cohort, we found that sub-AKI identified with isolated CysC increasing without satisfying SCr criteria might also be associated with worse outcomes including prolonged LOIS, LOHS or DMV, higher hospitalization expense. Meanwhile, the worst outcomes were demonstrated in the patients with AKI both detected by SCr and CysC. Previous study has shown that SCr is more associated with kidney functions when it was in a relative stable condition^23^. Therefore, in the process of developing AKI, the SCr might not reflect the fluctuating kidney function in a short time period. Due to its inherent characteristics, CysC was demonstrated to be much superior marker of AKI^24^ and with great accuracy to predict AKI in children^25^, which was reflected by the results of our study with worse outcomes detected in isolated CysC increase. In addition to CysC alone, combined SCr and CysC accurately predict the worst outcomes. Inconsistent with previous study, Zappitelli et al. reported the highest urine biomarkers concentration in patients with both SCr and CysC increasing^12^. Our study enrich evidence reflecting the use of combined SCr and CysC much related to worse clinical outcomes. This suggests that information from both SCr and CysC change may be useful in clinical care in which specificity of AKI diagnosis is most desirable.

In our clinical practice, serum CysC was routinely monitored for the perioperative patients. However, when aiming to diagnose AKI, physicians preferred the SCr criteria, which might result from their concerning of the diagnostic accuracy, threshold and agreement with SCr criteria. In our study, although agreement in the presence of any AKI was only moderate between SCr-AKI and CysC-AKI, agreement between identifying stage 2 CysC-AKI and SCr-AKI was very high. This may suggest that CysC rise may be more specific for denoting the presence of true and more severe acute renal tubular injury. However, about 70% of the patients with stage 1 SCr-AKI did not have an CysC-AKI. It is possible that patients classified as having SCr-AKI immediately after surgery did not have actual renal tubular injury but simply had immediate postoperative SCr rise due to creatinine production or secretion or metabolism.

Furthermore, our study assessed the predict accuracy of CysC at different time points for AKI in pediatric patients undergoing cardiac surgery. Our results showed that postoperative CysC higher than the cutoff value of 1.29 mg/dL might be used to identify greater risk of AKI in our patients setting. The cutoff value of relative variation of CysC or preoperative CysC might also have a predict value, while its diagnostic accuracy might be lower than isolated postoperative CysC. Due to its inversely correlation with age and body weight^26-28^, the absolute value of CysC might much directly predict AKI. Our single center cohort study with a large pediatric population provides adequate power for validating threshold though the generalizability of our findings remained concerning. Our data provide a valuable measure to refine the current definition of AKI and supply evidence that postoperative serum CysC at the cutoff of 1.29 mg/dL is not only helpful in the discrimination of AKI, but also indicative of AKI subtypes. Potential utility of serum CysC is underlined by the proof of a prognostic impact associated with AKI subtypes.

In contrast to the previous studies which focusing on the earlier diagnostic value of serum CysC for AKI, we also assessed the effect of CysC in distinguishing patients with higher risk for worse outcomes. Patients with both SCrAKI and CysC-AKI had longer LOIS, LOHS and DMV, higher hospitalization expense again supporting that the inclusion of CysC in determining if AKI is present increases association of AKI with outcomes.

Our study has some limitations. First, it was a single-center study, which led to a limited number of patients, and it is unclear whether our results can be generalized at this point. Second, we did not collect the required data to incorporate the urine output criteria for defining AKI and were unable to explore if SCr-AKI vs CysC-AKI definitions are associated with different urine output indexes. Third, the sample size of our study was inadequate to evaluate the association for 30-day mortality, resulting in reliability of our adjusted HR for 30-day mortality in our study. Forth, we calculated baseline eGFR using modified Schwartz formula, while it remains uncertain whether it is the best equation. Fifth, because the number of deaths with stage 2 and 3 AKI under each criterion in our study was too small, we were unable to verify the association between each severity of AKI and clinical outcomes. In addition, we did not compare CysC to other biomarkers of renal damage (e.g., NGAL, kidney injury molecule-1, or liver-type fatty acid-binding protein). Whether or not the combination of CysC and other biomarkers further improve the ability in defining AKI phenotypes needs to be explored.

## Conclusions

Our analysis indicates defining AKI with both CysC and SCr might more significantly affecting clinical outcome associations in pediatric patients undergoing cardiac surgery. Moreover, the serum CysC cutoff of 1.29mg/dL postoperatively is a valuable threshold for AKI risk assessment to define AKI subtypes. Further clinical trials are needed to explore whether timely and personalized intervention based on AKI subtypes would improve the clinical outcomes in pediatric patients undergoing cardiac surgery.

## Data Availability

The data that support the findings of this study are available from the corresponding author upon reasonable request.

## References

1. Kaddourah A, Basu RK, Bagshaw SM, Goldstein SL. Epidemiology of Acute Kidney Injury in Critically Ill Children and Young Adults. N Engl J Med. 2017;376:11–20. doi: 10.1056/NEJMoa1611391

2. Jetton JG, Boohaker LJ, Sethi SK, Wazir S, Rohatgi S, Soranno DE, Chishti AS, Woroniecki R, Mammen C, Swanson JR, et al. Incidence and outcomes of neonatal acute kidney injury (AWAKEN): a multicentre, multinational, observational cohort study. Lancet Child Adolesc Health. 2017;1:184–194. doi: 10.1016/s2352-4642(17)30069-x

3. Blinder JJ, Goldstein SL, Lee VV, Baycroft A, Fraser CD, Nelson D, Jefferies JL. Congenital heart surgery in infants: effects of acute kidney injury on outcomes. J Thorac Cardiovasc Surg. 2012;143:368–374. doi: 10.1016/j.jtcvs.2011.06.021

4. Li S, Krawczeski CD, Zappitelli M, Devarajan P, Thiessen-Philbrook H, Coca SG, Kim RW, Parikh CR. Incidence, risk factors, and outcomes of acute kidney injury after pediatric cardiac surgery: a prospective multicenter study. Crit Care Med. 2011;39:1493–1499. doi: 10.1097/CCM.0b013e31821201d3

5. Park SK, Hur M, Kim E, Kim WH, Park JB, Kim Y, Yang JH, Jun TG, Kim CS. Risk Factors for Acute Kidney Injury after Congenital Cardiac Surgery in Infants and Children: A Retrospective Observational Study. PLoS One. 2016;11:e0166328. doi: 10.1371/journal.pone.0166328

6. Ostermann M, Zarbock A, Goldstein S, Kashani K, Macedo E, Murugan R, Bell M, Forni L, Guzzi L, Joannidis M, et al. Recommendations on Acute Kidney Injury Biomarkers From the Acute Disease Quality Initiative Consensus Conference: A Consensus Statement. JAMA Netw Open. 2020;3:e2019209. doi: 10.1001/jamanetworkopen.2020.19209

7. Albert C, Haase M, Albert A, Zapf A, Braun-Dullaeus RC, Haase-Fielitz A. Biomarker-Guided Risk Assessment for Acute Kidney Injury: Time for Clinical Implementation? Ann Lab Med. 2021;41:1–15. doi: 10.3343/alm.2021.41.1.1

8. Haase M, Kellum JA, Ronco C. Subclinical AKI--an emerging syndrome with important consequences. Nat Rev Nephrol. 2012;8:735–739. doi: 10.1038/nrneph.2012.197

9. Moledina DG, Parikh CR. Phenotyping of Acute Kidney Injury: Beyond Serum Creatinine. Semin Nephrol. 2018;38:3–11. doi: 10.1016/j.semnephrol.2017.09.002

10. Prowle JR, Rosner MH. Have biomarkers failed in acute kidney injury? We are not sure. Intensive Care Med. 2017;43:890–892. doi: 10.1007/s00134-017-4763-7

11. Vanmassenhove J, Kielstein JT, Ostermann M. Have renal biomarkers failed in acute kidney injury? Yes. Intensive Care Med. 2017;43:883–886. doi: 10.1007/s00134-017-4759-3

12. Zappitelli M, Greenberg JH, Coca SG, Krawczeski CD, Li S, Thiessen-Philbrook HR, Bennett MR, Devarajan P, Parikh CR. Association of definition of acute kidney injury by cystatin C rise with biomarkers and clinical outcomes in children undergoing cardiac surgery. JAMA Pediatr. 2015;169:583–591. doi: 10.1001/jamapediatrics.2015.54

13. Musiał K. Current Concepts of Pediatric Acute Kidney Injury-Are We Ready to Translate Them into Everyday Practice? J Clin Med. 2021;10. doi: 10.3390/jcm10143113

14. Schwartz GJ, Schneider MF, Maier PS, Moxey-Mims M, Dharnidharka VR, Warady BA, Furth SL, Muñoz A. Improved equations estimating GFR in children with chronic kidney disease using an immunonephelometric determination of cystatin C. Kidney Int. 2012;82:445–453. doi: 10.1038/ki.2012.169

15. Finney H, Newman DJ, Thakkar H, Fell JM, Price CP. Reference ranges for plasma cystatin C and creatinine measurements in premature infants, neonates, and older children. Arch Dis Child. 2000;82:71–75. doi: 10.1136/adc.82.1.71

16. Wald R, Liangos O, Perianayagam MC, Kolyada A, Herget-Rosenthal S, Mazer CD, Jaber BL. Plasma cystatin C and acute kidney injury after cardiopulmonary bypass. Clin J Am Soc Nephrol. 2010;5:1373–1379. doi: 10.2215/cjn.06350909

17. Zappitelli M, Krawczeski CD, Devarajan P, Wang Z, Sint K, Thiessen-Philbrook H, Li S, Bennett MR, Ma Q, Shlipak MG, et al. Early postoperative serum cystatin C predicts severe acute kidney injury following pediatric cardiac surgery. Kidney Int. 2011;80:655–662. doi: 10.1038/ki.2011.123

18. Section 2: AKI Definition. Kidney Int Suppl (2011). 2012;2:19–36. doi: 10.1038/kisup.2011.32

19. O’Brien SM, Clarke DR, Jacobs JP, Jacobs ML, Lacour-Gayet FG, Pizarro C, Welke KF, Maruszewski B, Tobota Z, Miller WJ, et al. An empirically based tool for analyzing mortality associated with congenital heart surgery. J Thorac Cardiovasc Surg. 2009;138:1139–1153. doi: 10.1016/j.jtcvs.2009.03.071

20. Schwartz GJ, Muñoz A, Schneider MF, Mak RH, Kaskel F, Warady BA, Furth SL. New equations to estimate GFR in children with CKD. J Am Soc Nephrol. 2009;20:629–637. doi: 10.1681/asn.2008030287

21. Erratum regarding “Pulmonary Hypertension in CKD” (Am J Kidney Dis. 2013;61[4]:612-622). Am J Kidney Dis. 2015;65:524. doi: 10.1053/j.ajkd.2014.12.004

22. Soto K, Coelho S, Rodrigues B, Martins H, Frade F, Lopes S, Cunha L, Papoila AL, Devarajan P. Cystatin C as a marker of acute kidney injury in the emergency department. Clin J Am Soc Nephrol. 2010;5:1745–1754. doi: 10.2215/cjn.00690110

23. Rule AD, Bergstralh EJ, Slezak JM, Bergert J, Larson TS. Glomerular filtration rate estimated by cystatin C among different clinical presentations. Kidney Int. 2006;69:399–405. doi: 10.1038/sj.ki.5000073

24. Dharnidharka VR, Kwon C, Stevens G. Serum cystatin C is superior to serum creatinine as a marker of kidney function: a meta-analysis. Am J Kidney Dis. 2002;40:221–226. doi: 10.1053/ajkd.2002.34487

25. Nakhjavan-Shahraki B, Yousefifard M, Ataei N, Baikpour M, Ataei F, Bazargani B, Abbasi A, Ghelichkhani P, Javidilarijani F, Hosseini M. Accuracy of cystatin C in prediction of acute kidney injury in children; serum or urine levels: which one works better? A systematic review and meta-analysis. BMC Nephrol. 2017;18:120. doi: 10.1186/s12882-017-0539-0

26. Li Y, Fu C, Zhou X, Xiao Z, Zhu X, Jin M, Li X, Feng X. Urine interleukin-18 and cystatin-C as biomarkers of acute kidney injury in critically ill neonates. Pediatr Nephrol. 2012;27:851–860. doi: 10.1007/s00467-011-2072-x

27. Saeidi B, Koralkar R, Griffin RL, Halloran B, Ambalavanan N, Askenazi DJ. Impact of gestational age, sex, and postnatal age on urine biomarkers in premature neonates. Pediatr Nephrol. 2015;30:2037–2044. doi: 10.1007/s00467-015-3129-z

28. Barbati A, Cappuccini B, Aisa MC, Grasselli C, Zamarra M, Bini V, Bellomo G, Orlacchio A, Di Renzo GC. Increased Urinary Cystatin-C Levels Correlate with Reduced Renal Volumes in Neonates with Intrauterine Growth Restriction. Neonatology. 2016;109:154–160. doi: 10.1159/000441273

